## Supplementary material for "Silicosis, tuberculosis and silica exposure among artisanal and small-scale miners: A systematic review and modelling paper": S1 File

### S1 Search strategy

We searched PubMed, Web of Science, Scopus and Embase for English and French language studies published before the 24th March 2023. We included primary research that included current or previous small-scale miners or mines.

Embase

1.	(mining or mine*).mp.

2.	exp mining/

3.	(small-scale or smallscale or artisan* or informal or galamsey).mp.

4.	1 or 2

5.	3 and 4

6.	exp respiratory tract disease/

7.	exp lung function test/

8.	exp respiratory system/

9.	exp lung/

10.	exp lung disease/

11.	exp tuberculosis/

12.	exp silicosis/

13.	(tuberculosis or silicosis or respiratory or lung or pulmonary or chest).mp.

14.	6 or 7 or 8 or 9 or 10 or 11 or 12 or 13

15.	5 and 14

Medline

1.	(mining or mine*).mp.

2.	Coal Mining/ or Mining/

3.	(small-scale or smallscale or artisan* or informal or galamsey).mp.

4.	1 or 2

5.	3 and 4

6.	exp Respiratory Tract Diseases/

7.	exp Respiratory Function Tests/

8.	exp Respiratory System/

9.	exp Lung Diseases/

10.	exp Lung/

11.	exp Tuberculosis/

12.	exp Silicosis/

13.	(tuberculosis or silicosis or respiratory or lung or pulmonary or chest).mp.

14.	6 or 7 or 8 or 9 or 10 or 11 or 12 or 13

15.	5 and 14

Web of Science

((artisanal OR small-scale OR informal OR galamsey) AND (mine OR miner OR mining)) AND (respirat* or lung* or pulmonary or silic* or tuberculosis or chest) (All fields)

Scopus

TITLE-ABS-KEY ( ( ( ( artisanal OR "small-scale" OR "smallscale" OR "small scale" OR informal OR galamsey ) AND ( mine OR miner OR mining ) ) AND ( respirat* OR lung* OR pulmonary OR silic* OR tuberculosis OR chest ) ) )
