## Supplementary material for "Silicosis, tuberculosis and silica exposure among artisanal and small-scale miners: A systematic review and modelling paper": S3 File

### S3 Data and quality extraction template

**Data extraction template**

**General information**

- Study ID
- Title
- Lead author contact details
- In which country was the study conducted
- Notes

**Characteristics of included studies**

**Methods**

- Aim of study
- Study design
  - Cohort study
  - Cross sectional study
  - Clinical trial
  - Other
- Start date
- End date
- Study funding sources
- Possible conflicts of interest for study authors

**Participants**

- Population description
- Inclusion criteria
- Exclusion criteria
- Definition of silicosis
- Definition of TB
- Description of spirometry
- Description of exposure measurement
- Method of recruitment of participants
  - Clinic patients
  - Screening population
  - Voluntary
  - Other
- Total number of participants/samples
- Baseline Population Characteristics
  - Number included in study
  - Age
  - Gender (%)
  - Duraton of work exposure (yrs)
  - Smokers (%)
  - History of tuberculosis
  - HIV prevalence
- Outcomes
  - Shortness of breath at rest
  - Cough
  - Wheeze
  - FEV1
  - FVC
  - FEV1/FVC
  - Microbiologically confirmed TB (%)
  - Clinically confirmed TB (%)
  - Radiographically confirmed silicosis (%)
  - Self-reported silicosis (%)
  - Prevalence of other lung conditions (%)

### Quality assessment template

**JBI - 1. Was the sample frame appropriate to address the target population?**

This question relies upon knowledge of the broader characteristics of the population of interest and the geographical area. If the study is of women with breast cancer, knowledge of at least the characteristics, demographics and medical history is needed. The term “target population” should not be taken to infer every individual from everywhere or with similar disease or exposure characteristics. Instead, give consideration to specific population characteristics in the study, including age range, gender, morbidities, medications, and other potentially influential factors.

**JBI - 2. Were study participants recruited in an appropriate way?**

Studies may report random sampling from a population, and the methods section should report how sampling was performed. Random probabilistic sampling from a defined subset of the population (sample frame) should be employed in most cases, however, random probabilistic sampling is not needed when everyone in the sampling frame will be included/ analysed. For example, reporting on all the data from a good census is appropriate as a good census will identify everybody.

**JBI - 3. Was the sample size adequate?**

The larger the sample, the narrower will be the confidence interval around the prevalence estimate, making the results more precise. An adequate sample size is important to ensure good precision of the final estimate. Ideally we are looking for evidence that the authors conducted a sample size calculation to determine an adequate sample size. This will estimate how many subjects are needed to produce a reliable estimate of the measure(s) of interest. Sometimes, the study will be large enough (as in large national surveys) whereby a sample size calculation is not required. In these cases, sample size can be considered adequate.

**JBI - 4. Were the study subjects and setting described in detail?**

Certain diseases or conditions vary in prevalence across different geographic regions and populations (e.g. Women vs. Men, sociodemographic variables between countries). The study sample should be described in sufficient detail so that other researchers can determine if it is comparable to the population of interest to them.

**JBI - 5. Was data analysis conducted with sufficient coverage of the identified sample?**

Coverage/Selection bias can occur when not all subgroups of the identified sample respond at the same rate. For instance, you may have a very high response rate overall for your study, but the response rate for a certain subgroup (i.e. older adults) may be quite low

**JBI – 6. Were valid methods used for the identification of the condition?**

Here we are looking for measurement or classification bias. Many health problems are not easily diagnosed or defined and some measures may not be capable of including or excluding appropriate levels or stages of the health problem. If the outcomes were assessed based on existing definitions or diagnostic criteria, then the answer to this question is likely to be yes. If the outcomes were assessed using observer reported, or self-reported scales, the risk of over- or under-reporting is increased, and objectivity is compromised. Importantly, determine if the measurement tools used were validated instruments as this has a significant impact on outcome assessment validity.

**JBI - 7. Was the condition measured in a standard, reliable way for all participants?**

Considerable judgment is required to determine the presence of some health outcomes. Having established the validity of the outcome measurement instrument (see item 6 of this scale), it is important to establish how the measurement was conducted. Were those involved in collecting data trained or educated in the use of the instrument/s?

**JBI - 8. Was there appropriate statistical analysis?**

Importantly, the numerator and denominator should be clearly reported, and percentages should be given with confidence intervals. The methods section should be detailed enough for reviewers to identify the analytical technique used and how specific variables were measured.

**JBI - 9. Was the response rate adequate, and if not, was the low response rate managed appropriately?**

A large number of dropouts, refusals or “not founds” amongst selected subjects may diminish a study’s validity, as can a low response rates for survey studies. The authors should clearly discuss the response rate and any reasons for non-response and compare persons in the study to those not in the study, particularly with regards to their socio-demographic characteristics.
