## Supplementary material for "Silicosis, tuberculosis and silica exposure among artisanal and small-scale miners: A systematic review and modelling paper": S6 File

### S6 Detailed description of modelling parameters

**Formula 1:**

Logit(p_1_) = Baseline silicosis risk + (B_1_* Cumulative RCS exposure)

p_1_ = the probability of silicosis

**Baseline silicosis risk** = the risk of silicosis in the lowest risk group (in this case 0 mg/m^3^ – however as the intercept may not equal 0, it can be considered analogous to the risk in the 0-1 mg/m^3^ group and was set at 2/100 cases. Additionally, it represents the proportion of false positives (approximately 2%(1,2)) on X-ray when a miner has not yet been exposed. The only study, by Miller et al, to use logistic regression and report their intercept found a value of 0.014(3). That we used a slightly higher intercept allows for a more gradual coefficient, as this study reported an odds of association of approximately 1.5.

**B_1_** = the odds of silicosis per 1 mg/m^3^-years increase in cumulative exposure of RCS. A default value of 1.3 is used, however it is essential to consider this value in the context of the population of interest as it will vary with latency and, potentially, intensity. Although many studies have been performed of low exposure settings with extended latent periods for measurement (thus allowing all possible cases to occur), no summary estimate of this figure is available in the literature. In addition to the study quote above by Miller et al, logistic regression odds of approximately 1.43 (95% CI 1.23-1.66) and 3.00 (2.24-3.83) in low exposure settings(4,5). Using measures of relative risk calculated from cox proportional hazards and Poisson regression, which can approximate the odds in the low incidence settingand higher risks are reported. For iron and copper mines, the hazard ratio was 1.41 (1.22-1.17), for Tin mines it was 1.14 (1.11-1.17), while for diatomaceous earth miners, an increased relative risk of 4.35 (1.7-11.06) was seen in the 1-3 mg/m^3^ group compared to those < 1 mg/m^3^ (5–7). Other studies use Weibull or log-logistic distributions to model cumulative risk, which are difficult to directly compare to models of relative risk.

Current research of ASM populations is characterised by high exposures with short latency to measurement, both of which are likely to modify the dose-response. Although high intensity exposures are considered to increase the odds of silicosis(3,7,8), short latency will reduce the odds as not all potential cases are accrued. In summary, the true risk or odds of silicosis in a given ASM population is not know. It may be however that compared to chronic, long latency silicosis, the *shape* of the relationship is similar. The relationship between cumulative exposure and the probability of silicosis at different odds is visualised in supplementary figure 3(9)). Based knowledge of current prevalence and exposures from this systematic review, previous studies of low intensity exposure with extended latent periods and the prevalence estimates of our model (figure XA) we estimated that a default odds of 1.3 was appropriate. As noted in the discussion of the intercept, by utilising a slightly higher intercept, we are able to have a slightly lower odds compared to some studies above, which does not lead to such rapid increases in risk at the higher ranges of RCS.

**Cumulative RCS Exposure** = takes the standard units of the number of years spent at an average exposure (mg/m^3^-years). This is estimated using a quadratic transformation of a normal distribution, to simulate a positively skewed distribution that is more often observed(3,7,10). To allow choice of the mean, the square root of the desired mean is taken and a fixed standard deviation of 0.5 mg/m^3^-year used to create a normal distribution, which is then squared. A series of illustrative distributions are given in the supplementary figure X, which demonstrate that the skew decreases and the standard deviation increases as the mean RCS value becomes larger – both plausible features. The default desired mean was 4 mg/m3-years.

**Formula 2:**

Logit(p_2_) = Baseline annual TB incidence + (B_1_*RCS exposure) + (B_2_*silicosis) + (B_3_*HIV)

**p_2_** = the probability of tuberculosis

**Baseline tuberculosis incidence** = the annual TB incidence per 100,000 in the general population for the WHO Africa region is 208 per 100,000 persons per year(11). The baseline in this model is for miners who are not silica exposed and not HIV positive. An accurate estimate for this figure is not available, however we may expect a slightly higher rate than the general population, as miners are generally young men may have other predisposing risk factors, such as nutritional status or a lower socioeconomic status. A conservative default value for miners who have no silica exposure and are HIV negative would therefore remain at 200 per 100,000 years.

**B_1_** = the odds of TB per 1 mg/m^3^-years increase in cumulative exposure of RCS. A meta-analysed estimate of TB risk in those exposed to silica and those not exposed demonstrates a pooled relative risk of 1.92 (95% CI 1.36, 2.73)(12). In a single study used in this meta-analysis the relative risk per 1 mg/m^3^-years of dust exposure, following adjustment for silicosis, was 1.07 (95% CI 1.04, 1.10)(13). Thus the default odds was thus, conservatively, set at 1.05.

**Cumulative RCS Exposure** = as described in formula 1.

**B_2_** = the odds of TB in the presence of silicosis compared to those without silicosis. A recent meta-analysis estimates this as 4.01 (95% CI 2.88, 5.58)(12) with a high confidence of the association. The default odds was therefore set at 4.

**Silicosis** = an indicator variable (0/1) as determined by formula 1.

**B_3_** = the odds of TB in the presence of HIV infection compared to those without HIV infection. In a cohort of South African gold miners in the 1990’s, the incidence of TB was 4.1 (95% CI 1.5-10.9) times higher in those with HIV than those without(14). That it was not higher than this, despite incomplete access to antiretroviral treatment (ART) may be due to a healthy worker effect.
