## Supplementary material for "Silicosis, tuberculosis and silica exposure among artisanal and small-scale miners: A systematic review and modelling paper": S7 Table

### S7. Table Characteristics of studies reporting spirometry (n=3) estimates among ASM

| Author, year | Study country, period | Study design | Population | Sample size | Sampling method | Spirometry method | Age (years), gender | FEV1 | FVC | FEV1/FVC (%) |
| --- | --- | --- | --- | --- | --- | --- | --- | --- | --- | --- |
| Osim, 1999(1)* | Zimbabwe, no date described | Exposure-control | Above and below ground chrome ASM, chrome LSM and community controls | 54 ASM , 46 LSM and 50 community controls | Not described | Three attempts at spirometry were allowed. Measurements required to meet ATS criteria (1979) | ASM: 32.7 +/- 1.5  LSM: 33.3 +/- 1.2  Controls: 31.6 +/-1.5,  All male | ASM: 2.6L +/- 0.1L  LSM: 3.1L +/- 0.1L  Controls: 3.2L +/- 0.1L | ASM: 3.5L +/- 0.1L  LSM: 3.9L +/- 0.1L  Controls: 3.2L +/- 0.1L | ASM: 76.2% +/- 2.4%  LSM: 81.8% +/- 1.4%  Controls: 80.7% +/- 1.5% |
| Rajaee, 2017(2) | Ghana, 2011 | Cross-sectional | ASM current and ex-gold miners and local community controls | 57 ever miners, 14 never miners | Random sampling within stratified geographic clusters for Kejetia | Comprehensive and standardised methodology described. Excluded if criteria not met (159/172 of overall sample) | Not described | ASM: Male 88.7% (SD +/- 11.1). Female 87.4% (SD +/- 14.0)  Controls: Male 93.8% (SD +/- 20.2)  Female 88.8% (SD +/- 9.4) | ASM: Male 92.7% (SD +/- 10.0). Female 92.0% (SD +/- 11.2). Controls: Male 92.7% (SD +/- 20.7). Female 94.3% (SD +/- 10.8) | ASM: Male 92.9% (SD +/- 9.0). Female 95.1% (SD +/- 10.9)  Control: Male 102.1% (SD +/- 3.8)  Female 91.0% (SD +/- 4.5) |
| Kyaw, 2020(3) | Myanmar, 2020 | Cross-sectional | ASM underground gold miners and local community controls | 18 ASM, 11 controls | Randomly recruited from town; no specified method | Three attempts with best result used. If not successful repeated until success | ASM: 37.6 +/- 15.2, 66% male  Controls: 56.1 +/- 13.9, 55% male | ASM: 2.49L (IQR 2.06-3.35L),  81.5% (IQR 71.3–90.8%)  Control: 2.01L (IQR 1.71-2.51L),  83.0% (IQR 64.5–91.5%) | ASM: 2.89L (IQR 2.26-3.37L),  75.5% (IQR 64.5–86.8%)  Controls: 2.11L (1.77–2.59L), 77.0% (IQR 55.5–80.0%) | - |

* Similar smoking prevalence; ASM 9/54 (16%), LSM 7/46 (15%) and controls 7/50 (15%).

\# ASM: 8/18 (44%) smokers, controls: 2/11 (18%) smokers. Significant methodological issues with study
