## Supplementary material for "Silicosis, tuberculosis and silica exposure among artisanal and small-scale miners: A systematic review and modelling paper": S8 Table

### S8 Table. Characteristics of studies reporting respiratory symptoms (n=3) and smoking (n=8) or substance misuse (n=2) estimates among ASM

| Author, year | Study country, period | Study design | Population | Sample size | Sampling method | Age (years), gender | Symptoms ASM | Symptoms Controls | Smoking or substance misuse prevalence |
| --- | --- | --- | --- | --- | --- | --- | --- | --- | --- |
| Symptoms | | | | | | | | | |
| Leon-Kabamba, 2018(1) | Democratic Republic of Congo, 2016 | Exposure-control | Above ground coltan miners and government office workers | 441 (199 ASM, 242 community controls) | 199/247 miners from single mine, controls from local administrative building | ASM: 32.8 +/- SD 8.3 years  Controls: 33.9 +/- 9.3 years  No gender data | Shortness of breath: 55/199 (27.6%)  Morning cough: 95/199 (47.7%)  Wheeze at rest: 85/199 (42.7%) | Shortness of breath: 6/242 (2.5%)  Morning cough: 6/242 (2.5%)  Wheeze at rest: 6/242 (2.5%) | ASM: 115/199 (58%)  Controls: 22/242 (9.1%) |
| Ralph, 2018(2) | Cameroon, 2018 | Cross-sectional | Current or retired above and underground gold ASM | 273 (174 ASM and 99 controls) | Mix of stratified random and convenience sampling | ASM: mode group 26-35 years (37.9%), 132/174 (75.9%) male  Controls: mode group 36-50 years (42.4%), 57/99 (57.6%) male | Breathlessness: 6/174 (3.6%)  Persistent cough: 54/174 (31.0%) | Breathlessness: 0/99 (0%)  Persistent cough: 9/99 (9.1%) | ASM: 72/174 (41.4%) |
| Souza, 2021(3) | Brazil, 2017-2018 | Cross-sectional | Current underground precious stone ASM | 258 | Miners from 49/277 randomly selected mines | Mean 40 (SD +/- 16.0) years, all male | Dyspnoea: 29/258 (11.2%)  Cough: 65/258 (25.2%) | - | 46/258 (17.8%) |
| Smoking | | | | | | | | | |
| Osim, 1999(4) | Zimbabwe, no date described | Exposure-control | Above and below ground chrome ASM, chrome LSM and community controls | 54 ASM , 46 LSM and 50 community controls | Not described | ASM: 32.7 +/- 1.5  LSM: 33.3 +/- 1.2  Controls: 31.6 +/-1.5,  All male | - | - | ASM: 9/54 (16%)  LSM: 7/50 (14%)  Controls: 7/50 (15%) |
| Tse, 2007(5) | China, 1997-2001 | Cross-sectional | Retired underground gold ASM rock-drillers | 583 | Complete sample of rock-drillers | 24.4 +/- SD 6.7 years, All male | - | - | 115/583 (35.1%) |
| Souza, 2017(6) | Brazil, 2013-2014 | Cross-sectional | Current and retired underground precious stone ASM | 348 | Sequential sample of registered miners attending annual screening | Mean 40.1 (SD +/- 11.9) years, all male | - | - | 81/348 (23.3%) |
| Kyaw, 2020(7) | Myanmar, 2020 | Cross-sectional | ASM gold miners and local community controls | 18 ASM, 11 controls | Randomly recruited from town; no specified method | ASM: 37.6 +/- 15.2, 66% male  Controls: 56.1 +/- 13.9, 55% male | - | - | ASM: 8/18 (44%)  Controls: 2/11 (18%) |
| Mbuya, 2023(8) | Tanzania, 2019-2021 | Cross-sectional | Below ground gemstone ASM | 330 | 15 randomly sampled miners from 22 randomly chosen mines | Median 35.0 (IQR 30.0–44.0),  All male |  |  | 60/330 (18.2%) |
| Substance misuse | | | | | | | | | |
| Moyo, 2021(9) | Zimbabwe, 2020-2021 | Cross-sectional | Current artisanal gold miners | 514 | Miners attending TB outreach screening and hospital occupational health clinic | Mean 37.0 (SD +/- 12.7) years, 435/514 (85%) male | - | - | Marijuana use: 143/365 (28%) |
| Moyo, 2022(10) | Zimbabwe, 2020-2022 | Cross-sectional | Above or underground gold and chrome ASM | 3950 | Miners attending TB outreach screening and hospital occupational health clinic | Mean 35.5 (SD +/- 12.1) years, 3245/3950 (85%) male | - | - | "Substance use" 673/3098 (17.0%) |
