## Supplementary material for "Silicosis, tuberculosis and silica exposure among artisanal and small-scale miners: A systematic review and modelling paper": S10 Table

S8 Table 3. Estimates of silicosis prevalence (Table 3A) and tuberculosis incidence (Table 3B) at cumulative RCS distributions with a mean of 4, 8, 12 and 16 mg/m^3^-years based on simulation with a sample size of 10,000. Variables in the model are held constant, unless otherwise stated, at values of: a baseline silicosis prevalence of 2%, an increased odds of 1.3 of silicosis per 1 mg/m^3^-year increase in RCS exposure, a baseline TB incidence of 200 cases per 100,000 per year, an increased odds of 1.05 of TB per 1 mg/m^3^-year increase in RCS exposure, an increased odds of TB of 4x in those with silicosis, a randomly distributed baseline prevalence of HIV of 2% and an increased odds of TB of 2x in those with HIV.

Table 3A

| Mean cumulative RCS (mg/ mg/m^3^-year | Estimated prevalence of silicosis at levels of increasing silicosis risk per mg/m3-year increase in cumulative RCS | | |
| --- | --- | --- | --- |
|  | OR 1.2 | OR 1.3 | OR 1.5 |
| 4 | 4.2% | 6.5% | 12.6% |
| 8 | 9.0% | 17.4% | 38.6% |
| 12 | 17.7% | 35.8% | 67.8% |
| 16 | 30.6% | 57.1% | 87.5% |

Table 3B

| Mean cumulative RCS (mg/ mg/m^3^-year | Estimated annual incidence of TB per 100,000 at levels of increasing silicosis risk per mg/m3-year increase in cumulative RCS | | |
| --- | --- | --- | --- |
|  | OR 1.2 | OR 1.3 | OR 1.5 |
| 4 | 290 | 320 | 380 |
| 8 | 430 | 500 | 670 |
| 12 | 580 | 750 | 1130 |
| 16 | 860 | 1270 | 1650 |
