## Supplementary figures and images for "Silicosis, tuberculosis and silica exposure among artisanal and small-scale miners: A systematic review and modelling paper"

### S4 Figure

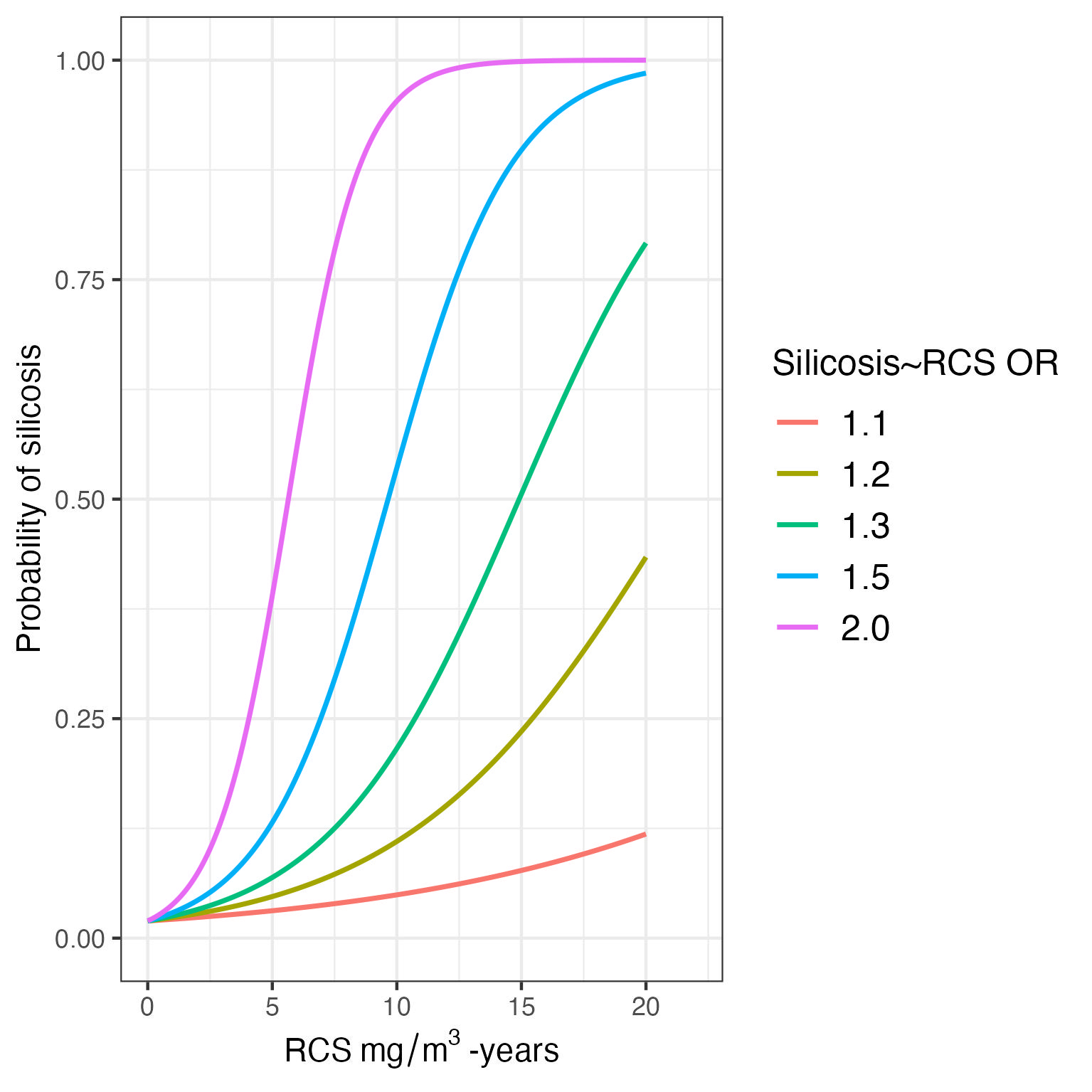

### S5 Figure

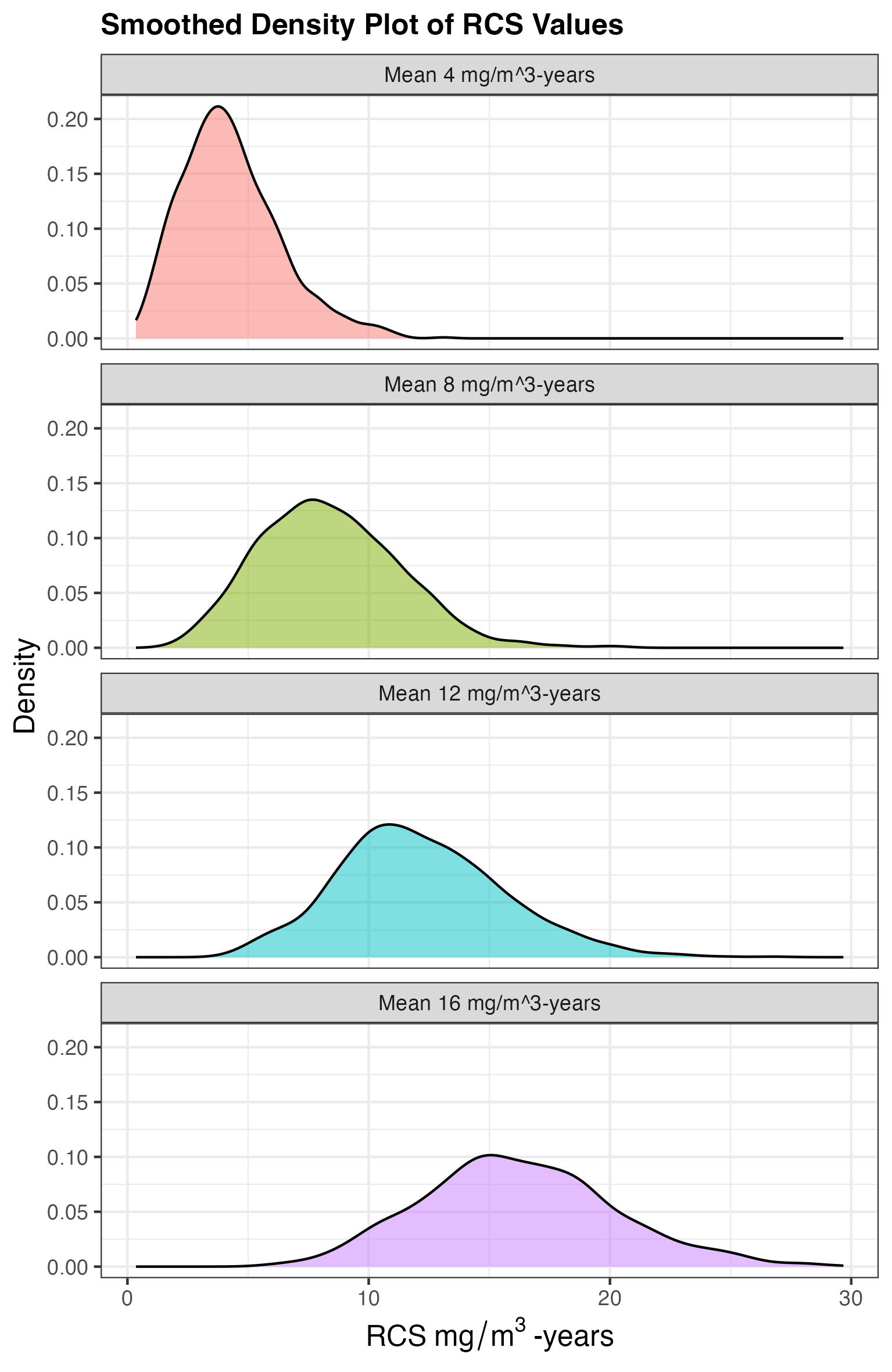

### S9 Figure

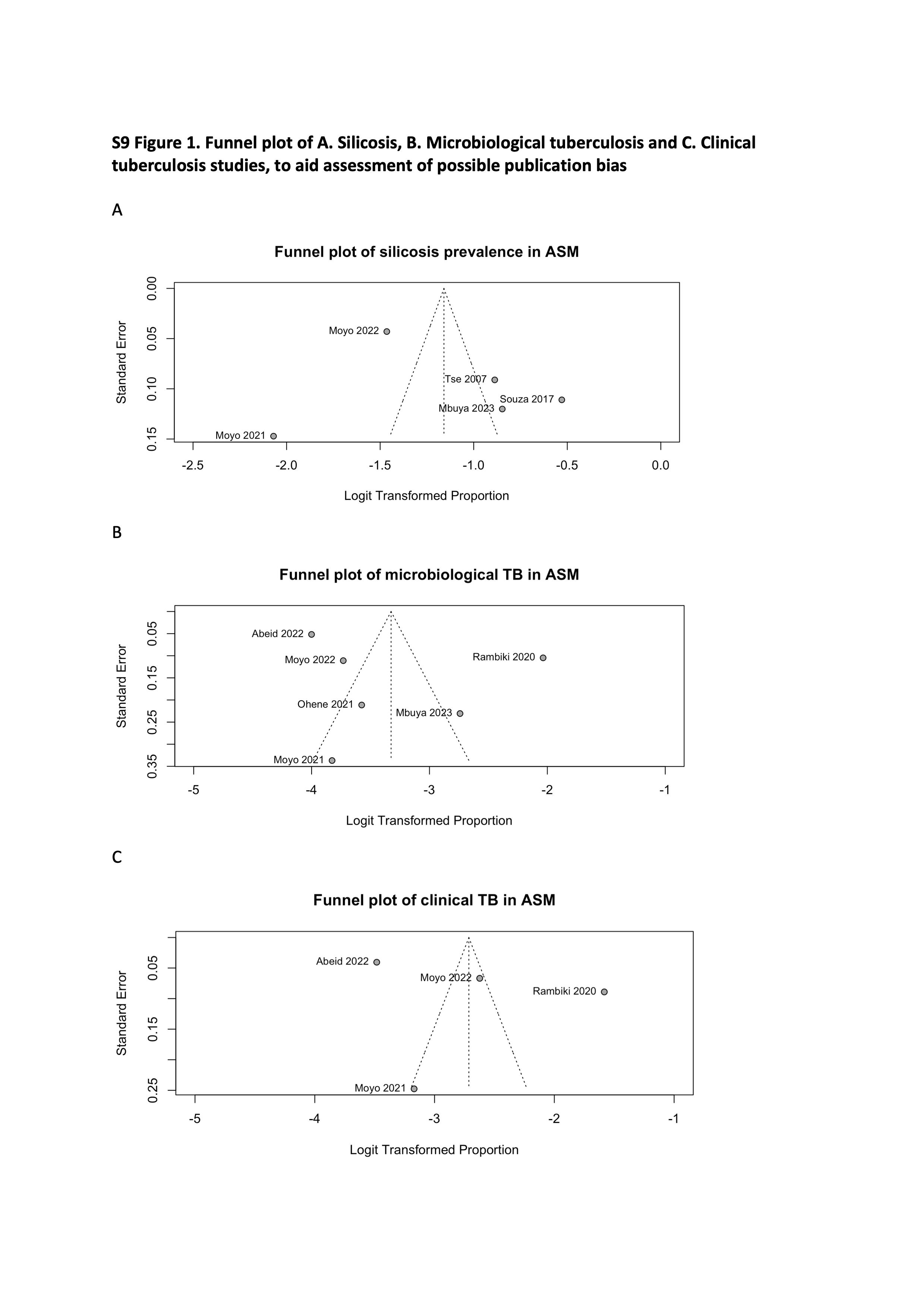
